# The seroprevalence of neutralizing antibodies against the adeno-associated virus capsids in Japanese hemophiliacs

**DOI:** 10.1101/2022.06.16.22276528

**Authors:** Yuji Kashiwakura, Nemekhbayar Baatartsogt, Shoji Yamazaki, Azusa Nagao, Kagehiro Amano, Nobuaki Suzuki, Tadashi Matsushita, Akihiro Sawada, Satoshi Higasa, Naoya Yamazaki, Teruhisa Fujii, Taemi Ogura, Hideyuki Takedani, Masashi Taki, Takeshi Matsumoto, Jun Yamanouchi, Michio Sakai, Masako Nishikawa, Yutaka Yatomi, Koji Yada, Keiji Nogami, Ryota Watano, Takafumi Hiramoto, Morisada Hayakawa, Nobuhiko Kamoshita, Akihiro Kume, Hiroaki Mizukami, Shizukiyo Ishikawa, Yoichi Sakata, Tsukasa Ohmori

**Author notes:** These authors contributed equally (Yuji Kashiwakura, Nemekhbayar Baatartsogt). **Correspondence should be addressed to T.O.**. Department of Biochemistry, Jichi Medical University School of Medicine, 3311-1 Yakushiji, Shimotsuke, Tochigi 329-0498, Japan.

## Abstract

Adeno-associated virus (AAV) vectors are promising modalities of gene therapy to address unmet medical needs. However, anti-AAV neutralizing antibodies (NAbs) hamper the vector-mediated therapeutic effect. Therefore, the NAb prevalence in the target population is vital in designing clinical trials with AAV vectors. Hence, updating the seroprevalence of anti-AAV NAbs, herein we analyzed sera from 100 healthy individuals and 216 hemophiliacs in Japan. In both groups, the overall seroprevalence against various AAV serotypes was 20%–30%, and the ratio of NAb-positive population increased with age. The seroprevalence did not differ between healthy participants and hemophiliacs and was not biased by the concomitant blood-borne viral infections. The high neutralizing activity, which strongly inhibits the transduction with all serotypes *in vitro*, was mostly found in the 60s or older age. The multivariate analysis suggested that “60s or older age” was the only independent factor related to the high titer of NAbs. Conversely, a large proportion of younger hemophiliacs was seronegative, rendering them eligible for AAV-mediated gene therapy in Japan. Compared with our previous study, the peak of seroprevalences has shifted to older populations, indicating natural AAV exposure in the elderly has occurred in their youth but not during the last decade.

## INTRODUCTION

Adeno-associated virus (AAV) vectors have been recently applied to *in vivo* gene therapy for various inherited diseases, including hemophilia. The expression of exogenous genes by systemic administration of an AAV vector is hindered by the presence of anti-AAV neutralizing antibodies (NAbs) related to previous AAV infection^1^. To date, most clinical trials for AAV-mediated hemophilia gene therapy have targeted only patients without NAbs against the AAV serotype used. Thus, it is essential to elucidate the seroprevalence of NAb before clinical trials.

Surveys for the seroprevalence of anti-AAV NAbs have shown broadly varying results, ranging from 2.2% to 96.6%^2–8^. Several factors appear to account for the prevalence of NAbs, including the geographical location. For example, high seroprevalence (∼96.6%) has been reported in India^4^, China^3^, and Korea^5^, compared with Europe^6^, the United States^7^, and Japan^8^. A recent global study of hemophilia A suggested the highest seropositivity to AAV5 in South Africa, Russia, Italy, and France compared with the United Kingdom, the United States, Germany, and Brazil^9^. Other factors, including ethnicity and age, are also known to affect the seroprevalence^3,7,9,10^. Previously, we reported that the seroprevalence in 40 years or older was much higher than those in younger populations in Japan^8^. Besides, seroprevalences differed among AAV serotypes. The seroprevalence against Clade B and C (AAV2 and AAV3B) that naturally infects humans displayed higher seropositivity than other serotypes, including Clade D and F (AAV8 and AAV9) ^2,7,11–13^.

This study aimed to determine the seroprevalence of NAbs against various AAV serotypes and NAb titers in hemophiliacs and healthy individuals in Japan, in order to estimate the number of patients with hemophilia eligible for future gene therapy in this cross-sectional study. We also explored the underling factors that influence seroprevalence and NAb titers. Our results will make it possible to discuss the mechanism of seroconversion and changes in NAb titers over time by comparing the results of this study with those of our previous survey conducted 10 years ago.

## RESULTS

### Study participants

We obtained 100 serum samples from each of 10 healthy men and women in the 20s, 30s, 40s, 50s, and 60s age range, and 227 hemophiliacs in Japan. Only the Japanese (not included other ethnicities) were enrolled in this study. Based on the exclusion criteria, 11 patients using emicizumab were excluded, resulting the total number of analyzed hemophiliacs as 216. Table 1 shows the profile of hemophiliacs and healthy volunteers in this study.

**Table 1.**
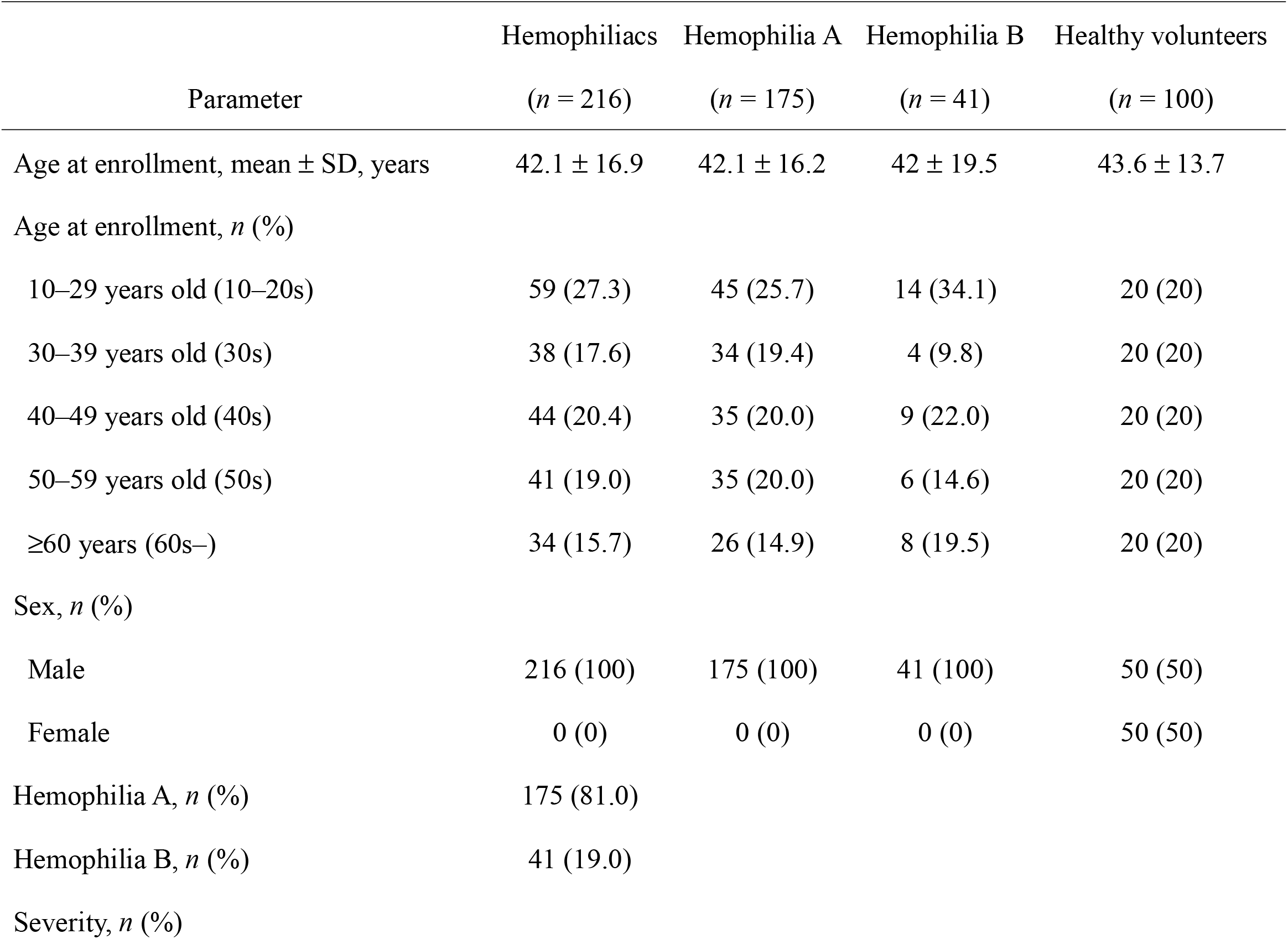
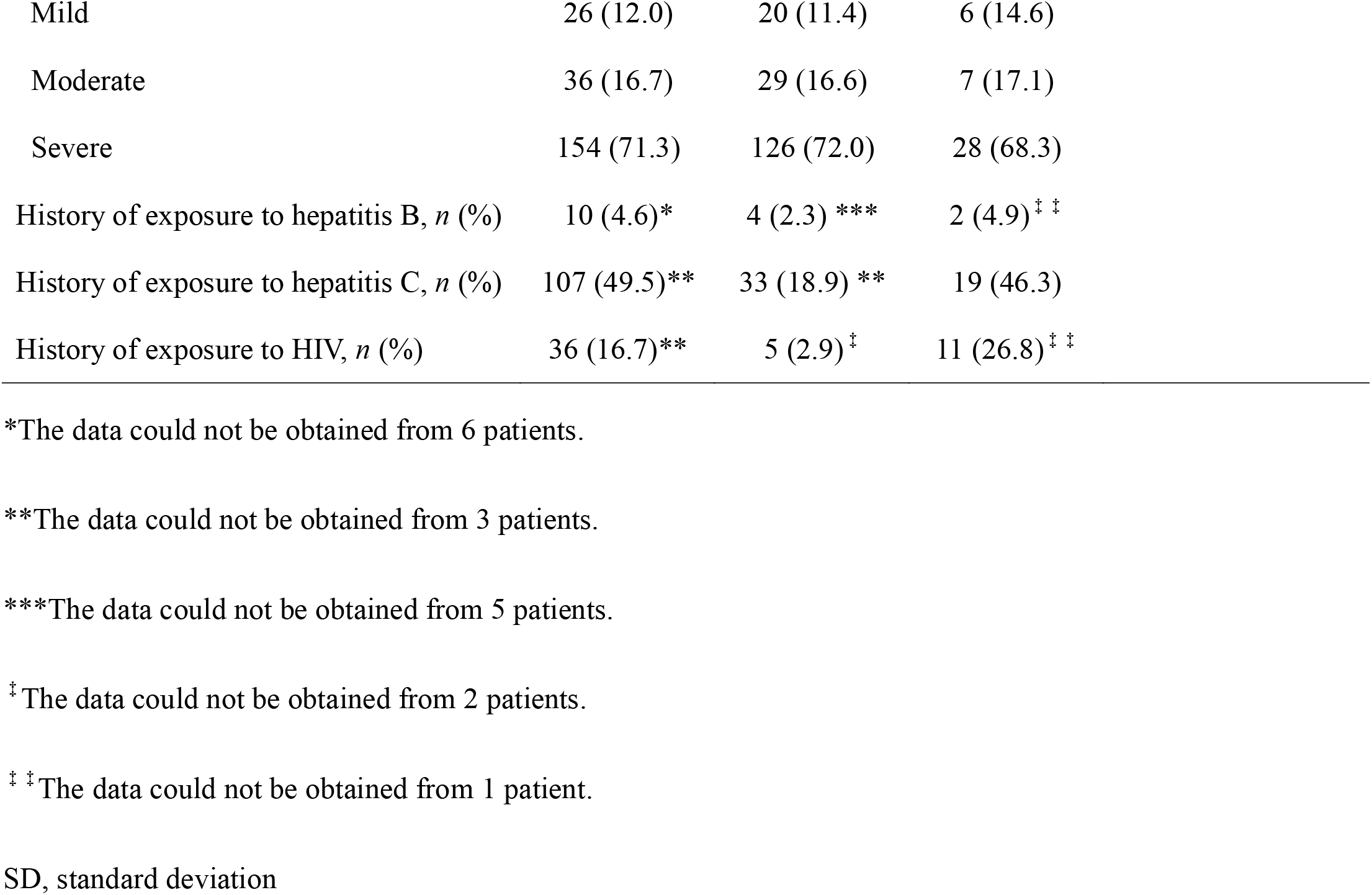
Characteristics of participants in this study

|  | Hemophiliacs | Hemophilia A | Hemophilia B | Healthy volunteers |
| --- | --- | --- | --- | --- |
| Parameter | (*n* = 216) | (*n* = 175) | (*n* = 41) | (*n* = 100) |
| Age at enrollment, mean ± SD, years | 42.1 ± 16.9 | 42.1 ± 16.2 | 42 ± 19.5 | 43.6 ± 13.7 |
| Age at enrollment, *n* (%) |  |  |  |  |
| 10–29 years old (10–20s) | 59 (27.3) | 45 (25.7) | 14 (34.1) | 20 (20) |
| 30–39 years old (30s) | 38 (17.6) | 34 (19.4) | 4 (9.8) | 20 (20) |
| 40–49 years old (40s) | 44 (20.4) | 35 (20.0) | 9 (22.0) | 20 (20) |
| 50–59 years old (50s) | 41 (19.0) | 35 (20.0) | 6 (14.6) | 20 (20) |
| $\geq 60$ years (60s–) | 34 (15.7) | 26 (14.9) | 8 (19.5) | 20 (20) |
| Sex, *n* (%) |  |  |  |  |
| Male | 216 (100) | 175 (100) | 41 (100) | 50 (50) |
| Female | 0 (0) | 0 (0) | 0 (0) | 50 (50) |
| Hemophilia A, *n* (%) | 175 (81.0) |  |  |  |
| Hemophilia B, *n* (%) | 41 (19.0) |  |  |  |
| Severity, *n* (%) |  |  |  |  |

|  |  |  |  |
| --- | --- | --- | --- |
| Mild | 26 (12.0) | 20 (11.4) | 6 (14.6) |
| Moderate | 36 (16.7) | 29 (16.6) | 7 (17.1) |
| Severe | 154 (71.3) | 126 (72.0) | 28 (68.3) |
| History of exposure to hepatitis B, *n* (%) | 10 (4.6)* | 4 (2.3) *** | 2 (4.9)<sup>‡ ‡</sup> |
| History of exposure to hepatitis C, *n* (%) | 107 (49.5)** | 33 (18.9) ** | 19 (46.3) |
| History of exposure to HIV, *n* (%) | 36 (16.7)** | 5 (2.9)<sup>‡</sup> | 11 (26.8)<sup>‡ ‡</sup> |

\*The data could not be obtained from 6 patients.
\*\*The data could not be obtained from 3 patients.
\*\*\*The data could not be obtained from 5 patients.
<sup>‡</sup> The data could not be obtained from 2 patients.
<sup>‡ ‡</sup> The data could not be obtained from 1 patient.
SD, standard deviation

### The seroprevalence of NAb against several AAV serotypes

We first assessed the seroprevalence of NAbs against various AAV serotypes in the healthy participants and hemophiliacs. The seroprevalence of NAbs to AAV1, AAV2, AAV3B, AAV5, AAV6, AAV7, AAV8, AAV9, and AAVrh10 was 20%–27% in healthy participants and 20.4%–29.2% in hemophiliacs (Figure 1A and B). No significant differences were observed in the frequency among serotypes (Figure 1A and B). In addition, the frequencies between patients and healthy participants were not significantly different, either (data not shown). When the cutoff NAb titer was increased from 1:1 to 1:10, 1:20, 1:50, and 1:100, the seroprevalence gradually decreased to 14%–20%, 11%–18%, 6%–17%, and 3%–16% in healthy participants, and 14.4%–21.3%, 11.6%–19.4%, 7.9%–17.1%, and 5.6%–14.4% in hemophiliacs, respectively (Figure 1E and 1F).

**Figure 1.**
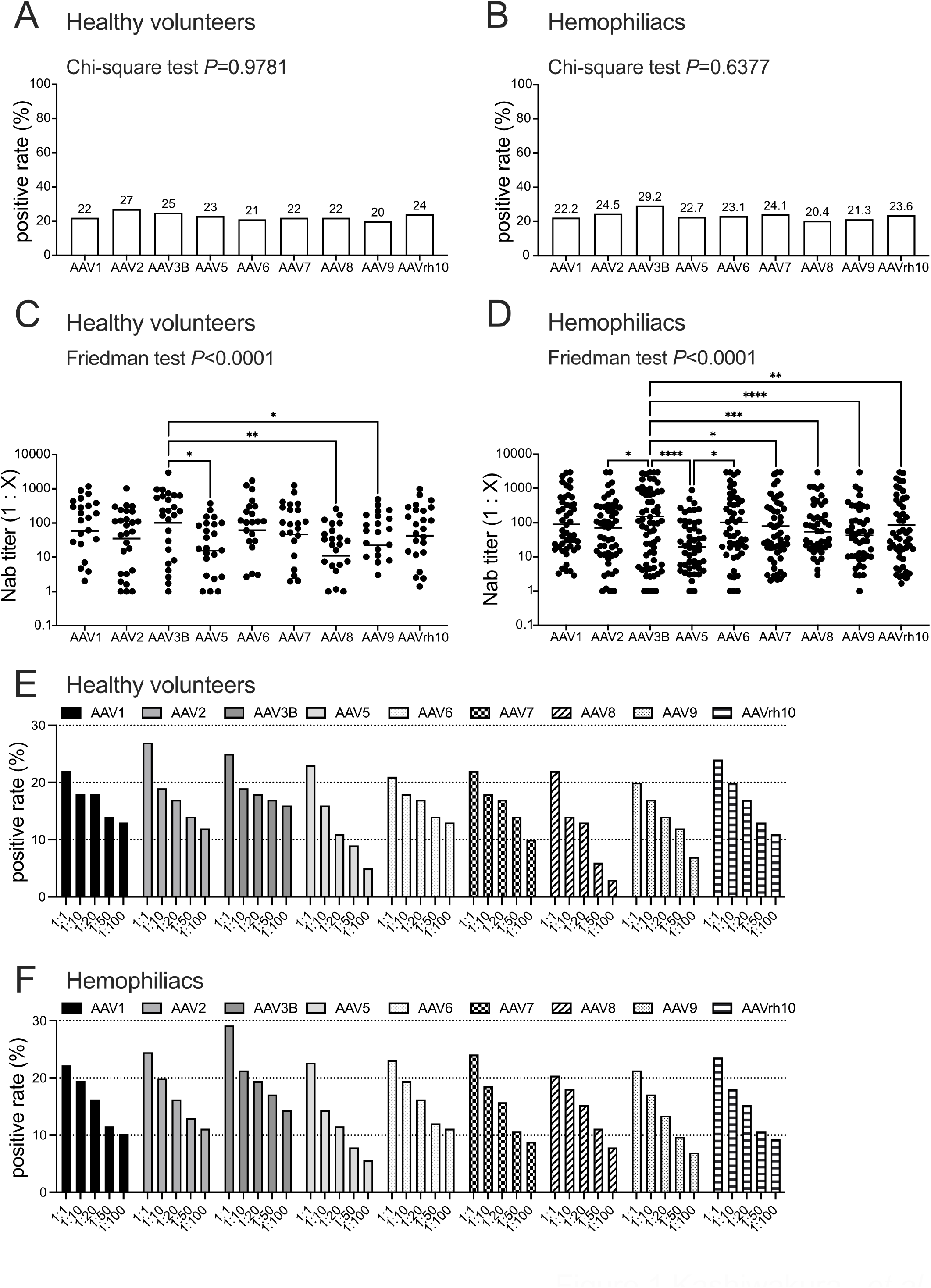
Seroprevalence and neutralizing titer of NAbs against each AAV serotype. (A and B) The seroprevalence of NAbs against each AAV in healthy volunteers (A) and hemophiliacs (B). The statistical difference among serotypes was analyzed using the *χ*^2^ test for trend. (C and D) The NAb titer against each AAV serotype in healthy volunteers (C) and hemophiliacs (D). (E and F) The seroprevalence of NAbs against each AAV in healthy volunteers (E) and hemophiliacs (F) at several thresholds. A positive rate is shown by each threshold of serum dilution factor (1:1, 1:10, 1:20, 1:50, and 1:100). Statistical differences among serotypes were analyzed using the Friedman test with the posthoc multiple comparisons test. \**P* < 0.05, \*\**P* < 0.01, \*\*\**P* < 0.001, and \*\*\*\**P* < 0.0001. AAV, adeno-associated virus; NAbs, neutralizing antibodies; n.s., not significant.

Then, we examined the NAb titers against each serotype (Figure 1C and D). The NAb titers against AAV3B were higher than those with AAV5, AAV8, and AAV9 in healthy participants, and higher than those against all serotypes, except AAV1 and AAV6, in hemophiliacs (Figure 1C and D). We observed no significant difference between healthy participants and patients in NAb titer of every AAV serotype (data not shown).

### The influence of sex, age, type and severities of hemophilia, and viral infections on the seroprevalence of NAb

Next, seroprevalence in males and females in healthy participants was compared. No difference was noted in seropositivity and NAb titers between men and women (data not shown). We further examined the impact of age on the seroprevalence and NAb titer (Figure 2). The seroprevalence of NAb was relatively low in the 20s, 30s, 40s, and 50s, and we observed no clear difference in the prevalence of each serotype when classified by age in these populations. Although the prevalence of AAV6 was 0% in 20s of healthy participants, there was no statistical difference of the prevalence between AAV6 and other serotypes (data not shown). In contrast, the seroprevalence was significantly higher in the 60s or older age than younger subjects (Figure 2, Figure S1 and S2; Table S1). No difference was observed in the prevalence between healthy participants and hemophiliacs by age group (data not shown). Furthermore, a comparison of the NAb titers between healthy participants and hemophiliacs revealed a small but significant difference between AAV8 NAbs in the 60s (Table S2).

**Figure 2.**
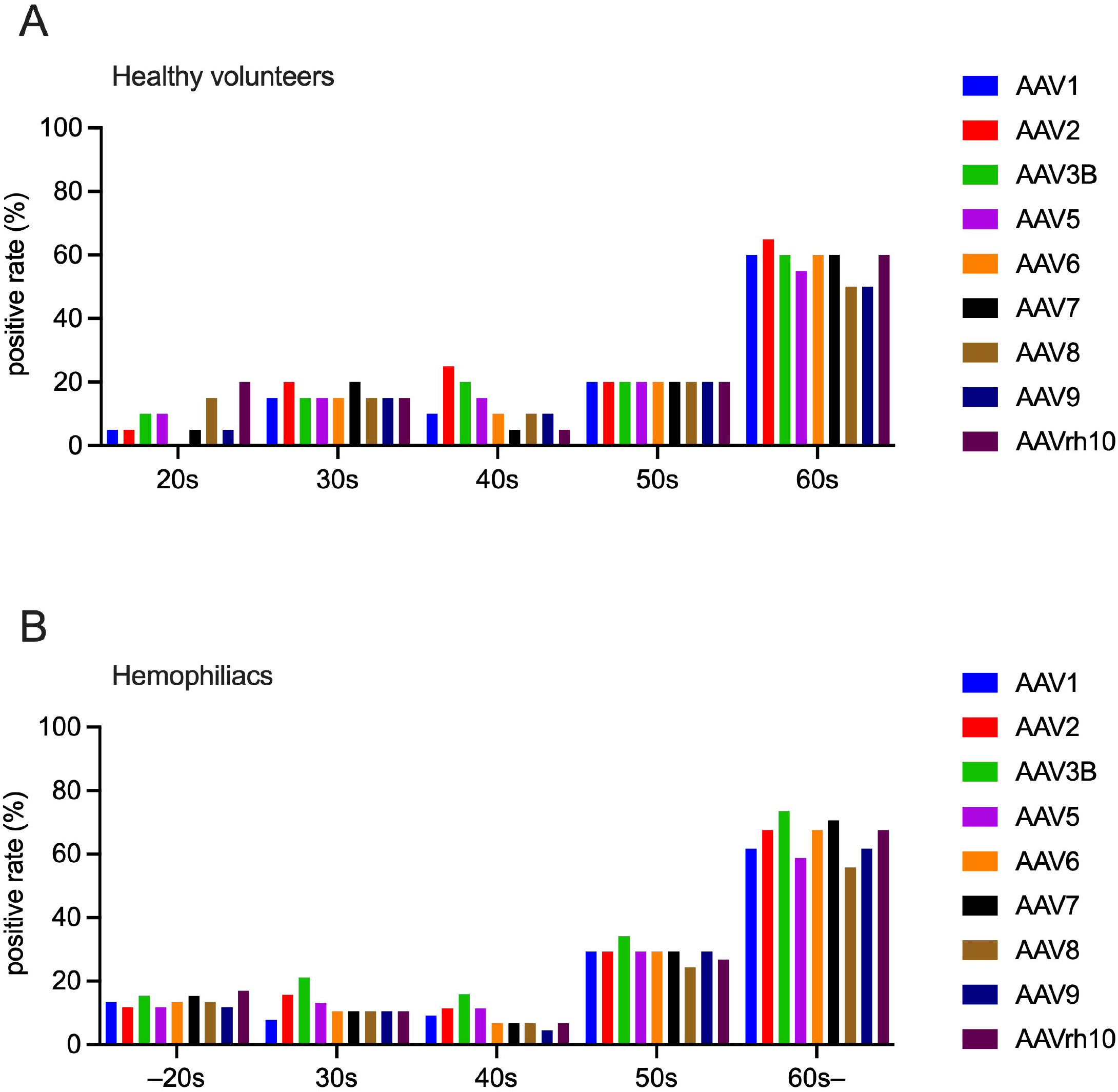
Differences in the seroprevalence of AAV NAbs among age groups. The seroprevalence of NAbs against each AAV of each age group (10–20s, 30s, 40s, 50s, 60s, or older) in healthy volunteers (A) and hemophiliacs (B). AAV, adeno-associated virus; NAbs, neutralizing antibodies.

In addition, we compared the seroprevalence and NAb titer between the type of hemophilia and the severity. No age difference was observed between hemophilia A and B (Figure S3A). The mean NAb prevalence was higher in hemophilia B patients than in hemophilia A, although the difference was not statistically significant (Figure S3B). However, the NAb titers were markedly higher in hemophilia B (Figure S3C). The severity of hemophilia did not influence the seroprevalence and NAb titer (data not shown).

Then, we determined whether the previous infection of hepatitis C virus (HCV), hepatitis B virus (HBV), and human immunodeficiency virus (HIV) affected anti-AAV NAbs (data not shown). The age of patients with a history of HCV and HIV, but not HBV, was markedly older than those without infections (data not shown). In addition, the concomitant infection of HCV, HBV, and HIV did not affect the relevance of NAbs. Besides, the NAb titers of several AAVs were significantly higher in HCV-positive participants. However, we observed no significant differences for those aged <50 years (data not shown). We further compared the prevalence between eastern and western region in Japan, and did not find any difference between two geographical regions (data not shown).

### Cross-reaction of NAbs with different serotypes

We examined the cross-reactivity of NAbs against multiple AAV serotypes. As shown in Figure 3, one-half of the NAbs reacted with all the AAV serotypes examined, which, in turn, were mostly found in participants aged 50 years or older, irrespective of the presence of hemophilia (Figure 3A and B). In contrast, NAbs reacting with only a single AAV serotype were found mostly in the 20–40 age groups (Figure 3A and B). The number of subjects without NAbs against any AAV serotypes in healthy volunteers and hemophilia patients was 63 (63%, 63/100) and 149 (69%, 149/216), respectively. Antibody titers were strikingly high when NAbs reacted with all serotypes (Figure 3C and D). Next, a multivariate logistic regression analysis was performed to identify participants with NAb cross-reacted with all serotypes. We included the variables of age (10–50s age group vs. 60 or older group), type of hemophilia (A vs. B), severity (mild and moderate vs. severe), HCV (negative vs. positive), HBV (negative vs. positive), and HIV (negative vs. positive). The age (60 years or older) was the only independent variable to determine the participants with NAbs reacted with multiple AAV serotypes (Table 2).

**Table 2.** Multivariate logistic regression analysis to examine neutralizing antibody against the multiple AAV serotypes

| Variables | Reference [0] | $\beta$ | Standard error | $P$ (Prob > $\chi^2$) | Odds ratio (95% CI) |
| --- | --- | --- | --- | --- | --- |
| Age | 10–50s | −1.0868764 | 0.2438719 | <0.0001* | 8.79 (3.45–23.66) |
| Type | Hemophilia A | −0.2478077 | 0.2495874 | 0.3208 | 1.64 (0.60–4.29) |
| Severity | Mild and moderate | −0.1175052 | 0.2347134 | 0.6166 | 1.26 (0.52–3.32) |
| HBV | None | −0.0136041 | 0.4602926 | 0.9764 | 1.03 (0.13–5.33) |
| HCV | None | −0.2251832 | 0.2490041 | 0.3658 | 1.57 (0.59–4.22) |
| HIV | None | 0.07752898 | 0.2903081 | 0.7894 | 0.85 (0.26–2.60) |

AAV, adeno-associated virus; CI, confidence interval; HBV, hepatitis B virus; HCV, hepatitis C virus; HIV, human immunodeficiency virus

**Figure 3.**
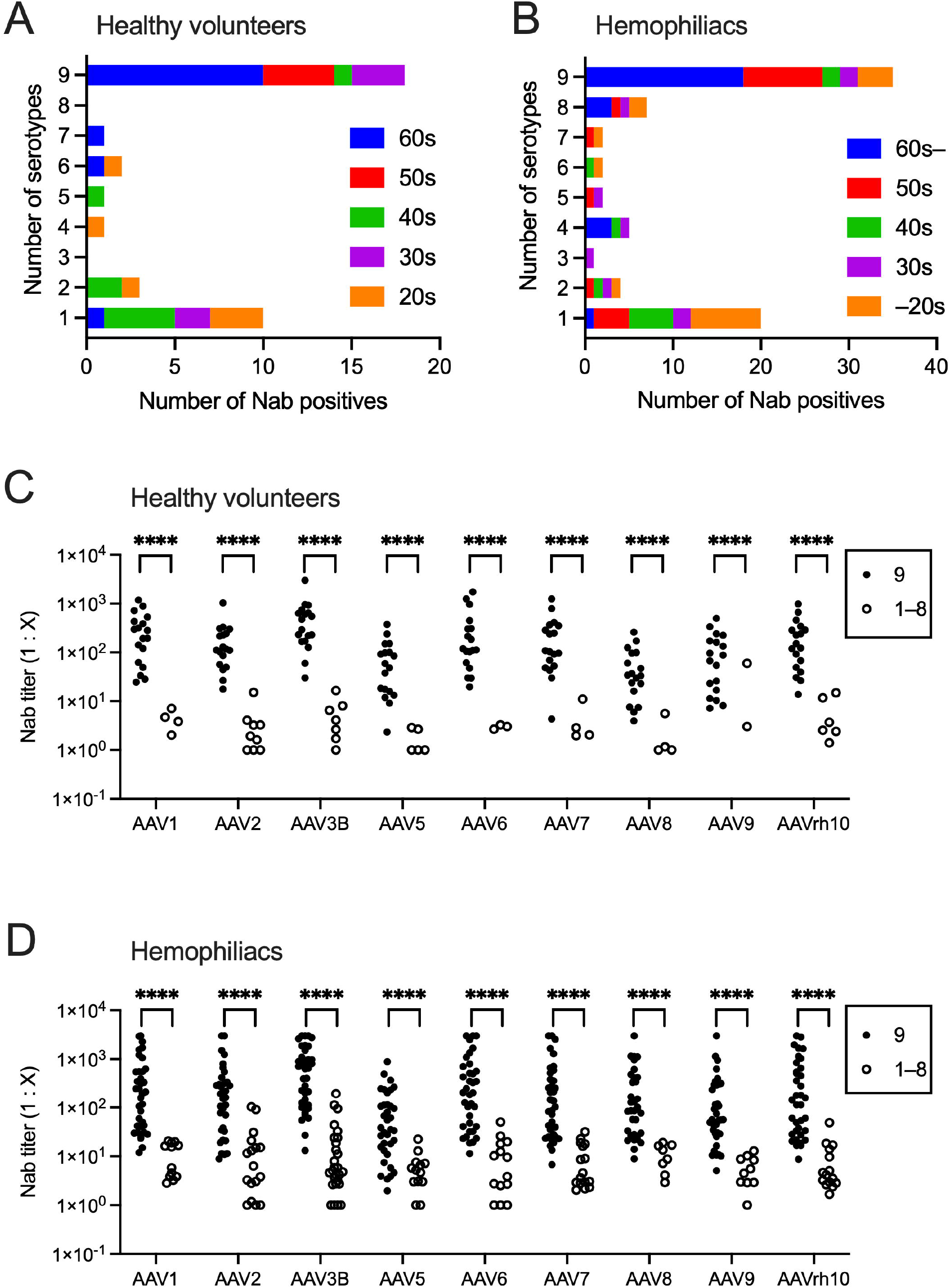
Cross-reaction of NAb with different AAV serotypes. (A and B) The number of serotypes inhibited by each positive serum in healthy volunteers (A) and hemophiliacs (B). The bars are shown to identify the proportion of age groups (10–20s, 30s, 40s, 50s, 60s, or older). (C and D) The comparison of NAb titers between the cases with antibodies to all serotypes (9 positives) and others (1–8 positives) in healthy volunteers (C) and hemophiliacs (D). Between-group differences were determined by the Mann–Whitney *U* test. \**P* < 0.05, \*\**P* < 0.01, \*\*\**P* < 0.001, and \*\*\*\**P* < 0.0001. AAV, adeno-associated virus; NAbs, neutralizing antibodies; n.s., not significant.

### Correlation of NAb titers among serotypes

We next examined the correlation of NAb titers among serotypes. The NAb titers of all the serotypes exhibited strong cross-correlations with each other (*P* < 0.00001 by Spearman’s rank correlation coefficient; Figure 4). Figure 4A shows the topology based on the similarity of the capsid sequences of various serotypes. The correlation of NAb titers was high between AAV1 and AAV6 (Figure 4B and C). Conversely, the correlation between NAbs to AAV5, the most distant serotype as per the topology, and those to the other serotypes was relatively low (Figure 4B and C). The factor analysis to identify latent variables and determine the variability of NAb titers among serotypes suggested that only one factor explained 89.9% of the variation (Figure 4D).

**Figure 4.**
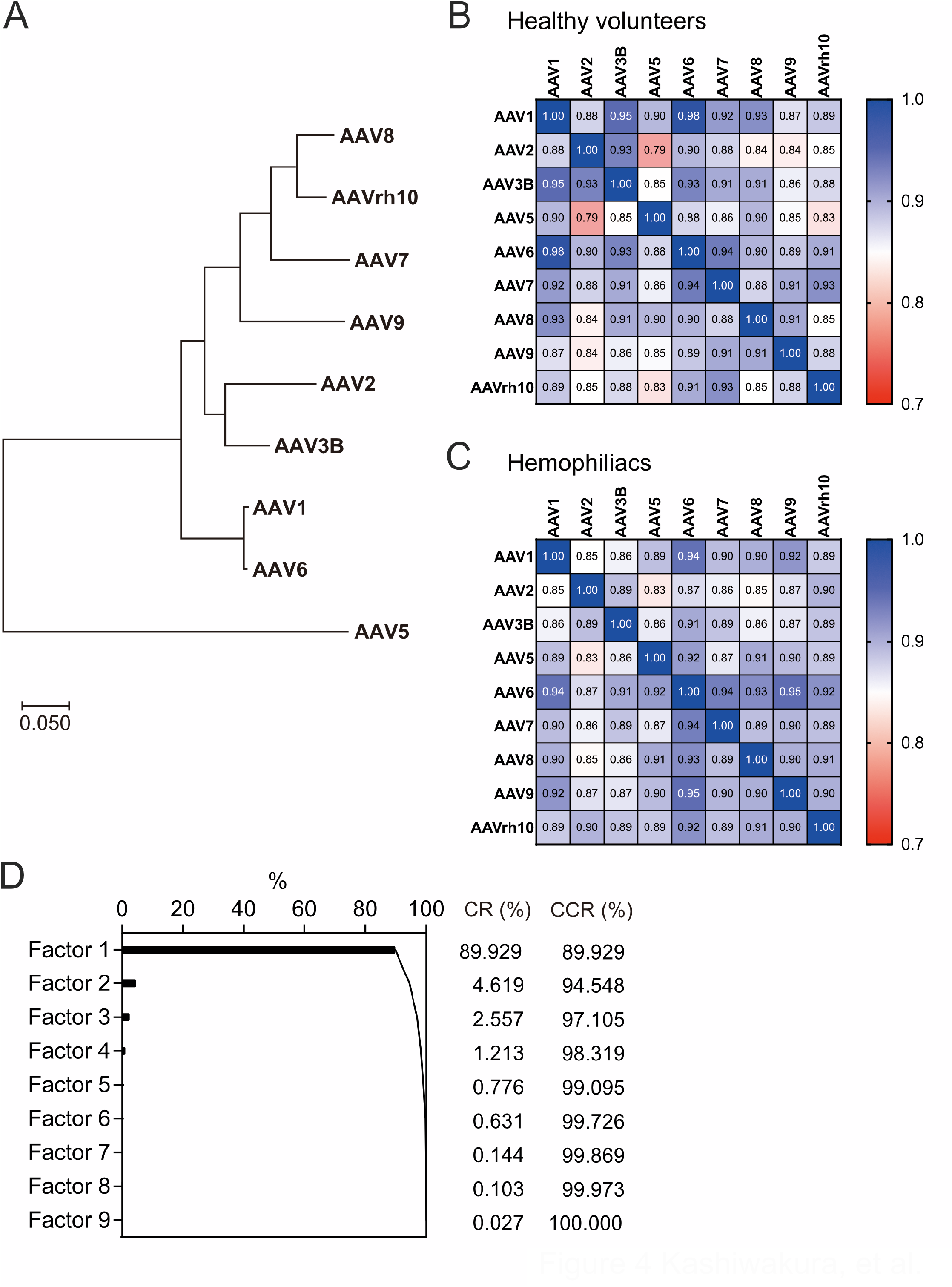
Correlation between NAb titers against each AAV serotype. (A) The topology is based on the similarity of the capsid sequences of various AAV serotypes. (B and C) The correlation between NAb titers against each AAV serotype in healthy volunteers (B) and hemophiliacs (C). The correlation coefficient was measured using the Spearman rank correlation coefficient. (D) The factor analysis to identify latent variables determines the variability of NAb titers among serotypes. AAV, adeno-associated virus; CR, contribution ratio; CCR, cumulative contribution ratio; NAbs, neutralizing antibody.

### Comparison with other ethnicity

Finally, we compared the abovementioned results with the NAb seroprevalence in other populations. To identify ethnical differences, sera from 10 Caucasians and 10 African–Americans aged 19–29 years in the United States were purchased. We selected subjects with hemophilia aged 19−29 years and healthy subjects aged 20s (same age group as US subjects) for comparison with healthy US subjects. The prevalence in Caucasians was 0%–20%, not much different from those in young Japanese (Figure 5). However, most African–Americans possessed NAb against AAVs (Figure 5). The prevalence of African–Americans, but not Caucasians, was significantly higher than those of age-matched Japanese hemophilia patient group and healthy subjects (*P*<0.0001 in AAV1, AAV2, AAV3B, AAV6, AAV7, AAV8, and AAV9; *P*=0.0004 in AAV5; *P*=0.0008 in AAVrh10 by chi-square test).

**Figure 5.**
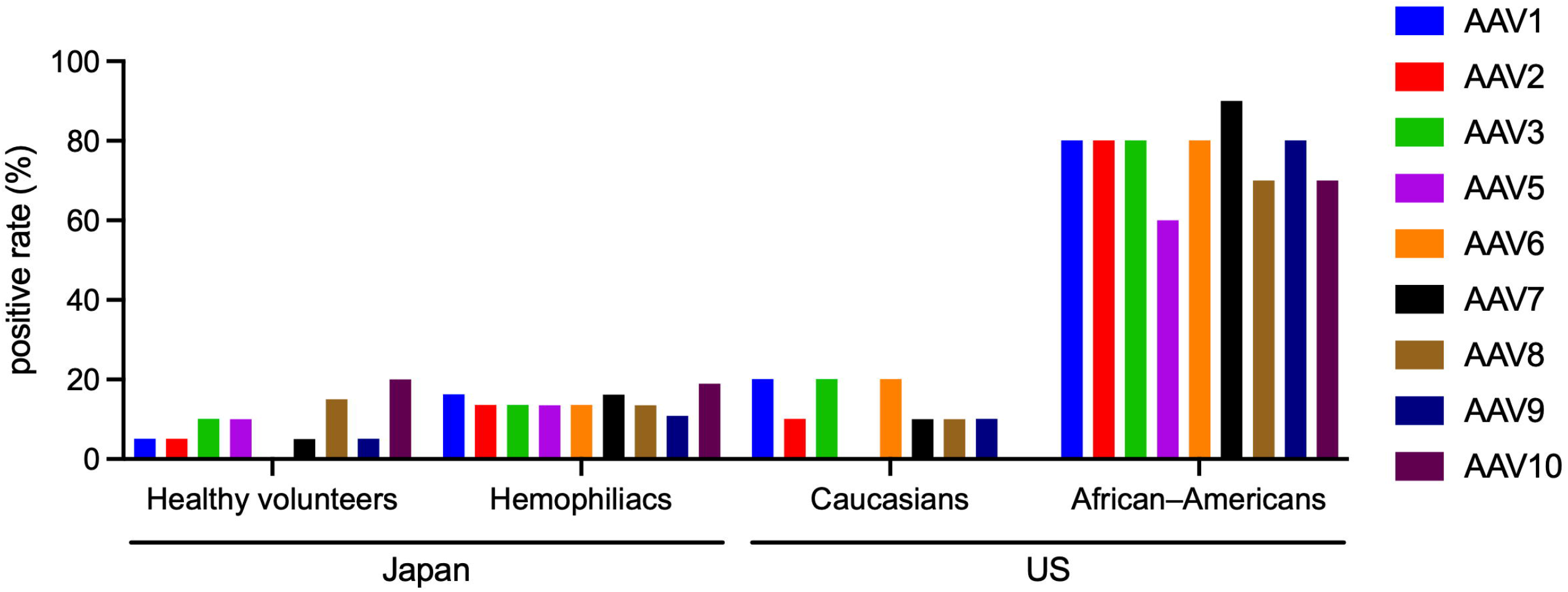
Differences in the seroprevalence of AAV NAbs among ethnicities at young ages. The seroprevalence of NAbs against each AAV in healthy volunteers (20s, *n* = 20) and hemophiliacs (aged 19–29 years, selected from the hemophiliacs, *n* =37) in Japan, Caucasians (*n* = 10), and African–Americans (*n* = 10). AAV, adeno-associated virus; NAbs, neutralizing antibodies.

## DISCUSSION

Gene therapy by intravenous AAV vector administration has been clinically applied to patients with hemophilia and neurological disorders^14^. In Japan, onasemnogene abeparvovec was approved as the first AAV gene therapy drug in 2020, for children aged <2 years with spinal muscular atrophy^15^. As the NAbs against AAV inhibit the efficacy of gene therapy with intravenous injection of AAV vectors, it is imperative to know the prevalence of NAbs in the region where the gene therapy is to be introduced before the clinical trial. This study measured the latest prevalence of NAbs and NAb titers against various AAV serotypes in Japan and demonstrated that the current seroprevalence of NAbs in Japan was one of the lowest among other countries. According to the recent survey in Japan^16^, there are 6,909 total hemophilia patients (5,657 with hemophilia A and 1,252 with hemophilia B) in Japan. Based on our survey of NAb prevalence, we estimated that 4,892−5,500 hemophilia (4,005−4,503 hemophilia A, and 886−997 hemophilia B) would be eligible for gene therapy using AAV vectors. Thus, many hemophiliacs, and possibly other target diseases of gene therapy, in Japan, especially in younger age groups, would benefit from AAV-mediated gene therapy.

In this study, the only demographical determining factor of the NAb prevalence was age, and the cutoff age in the Japanese was 60 years. Factor analysis also suggested that age was the only factor to determine NAb titer. Although several studies have consistently reported the increase in seropositivity according to age^9,10,17^, no survey has presented a clear distinction from certain age group(s). Moreover, when one obtains AAV NAb though natural infection remains unclear. A study of 4-year-follow-up reported that the seroconversion occurred during early childhood for all serotypes^13^. Over the 6 months observation, very few participants seroconverted and no geographic trend was noted^9^. In addition, the NAb titers remained unchanged during the 6-year-follow-up in Duchenne muscular dystrophy^18^. Compared with our survey of Japanese participants conducted 10 years ago^8^, the positive rate of anti-AAV antibodies among 41–50 years has clearly declined from 80% to 20%, and the peak of prevalence has clearly shifted to 60 years and older. These findings indicated that endemic AAV infection leading to a demographical seroconversion has not occurred in the past decade. Remarkably, the prevalence of AAV NAbs is similar to that of the hepatitis A virus (HAV)^19^. A study examining 2430 sera from 12 Japanese prefectures in 2003 (17 years ago from this study) reported a significant seropositivity of anti-HAV antibody in 45 years or older^19^. As a result of the recent disappearance of endemic infection, the age-specific seroprevalence of anti-HAV antibody had directly shifted to the right (older ages) over the past 30 years^19^. HAV causes acute hepatitis in humans and its primary transmission occurs via ingestion of food or water contaminated with HAV. The enhancement of sanitary conditions has decreased the incidence of hepatitis A^19^. While respiratory infection is thought to be the primary route of infection for AAV^20^, adenoviruses can be transmitted through a variety of routes, including droplet, contact, and fecal-oral transmission^21^. It is conceivable that the prevalence of NAbs may increase in the environments where the opportunity for helper virus infection is high. Perhaps, past (especially 50 years ago or more) sanitary environmental factors in Japan might influence AAV infections in the elderly during their past childhood.

The NAb titer of the elderly participants tended to be high and reacted to multiple AAV serotypes, whereas the NAb titer of younger participants tended to exhibit a low titer that reacted with one or a few AAV serotype(s). Of note, NAbs for AAV show cross-reactivity against multiple AAV serotypes^17,22^. Indeed, a longitudinal observational study of chimpanzees over 10 years indicated that a single natural infection with AAV induced a broadly cross-reactive NAb response to multiple AAV serotypes^23^. Natural AAV has a unique biphasic lifecycle, including a productive and latent phase, and natural AAV2 (not AAV vector) was inserted into the host genome at *AAVS1* locus in the latent phase^24^. Reportedly, the latent AAV infection elicited NAbs but no cell-mediated immune responses in a nonhuman primate model^25^. The subsequent infection of helper viruses such as herpesvirus and adenovirus reactivates latent AAV, leading to proliferation and release of a large amount of AAV^25^. Once AAV is integrated into the host chromosome, repeated helper virus infection may facilitate the AAV proliferation and result in the high titer of NAbs against AAV. This is a similar situation to the observation that multiple doses of the SARS-CoV-2 vaccine increase NAb levels in strains with different antigenic properties^26^. Thus, repeated exposure to the helper virus in a latent infection in youth could be accountable for the high titer of NAbs reacted with multiple AAV serotypes in elderly participants.

This study suggests that the NAbs titers were consistently higher in patients with hemophilia B than in patients with hemophilia A. We did not clarify the actual reason for higher NAb titers in patients with hemophilia B, although a previous study suggested that using a plasma-derived factor concentrate correlated with the seroprevalence of AAV8 NAb^27^. We could not obtain the detailed history of previous treatments with plasma-derived products in individual cases, but the introduction of recombinant factor concentrates for hemophilia B was considerably delayed in Japan. Recombinant factor VIII product was approved in 1993, while recombinant factor IX was not available until 2010. Per the Nationwide Survey on Coagulation Disorders 2015^28^, 40.1% of patients with hemophilia B used plasma-derived coagulation factors (252/629 patients, including bypass reagents), while 15.5% of those with hemophilia A (439/2840 patients) in 2015. Perhaps, the delay in the widespread use of recombinant preparations could correlate with the high titer of AAV in hemophilia B. AAV is a small single-strand DNA virus belonging to the parvovirus family. Parvovirus cannot be efficiently eliminated through viral inactivation of plasma-derived factor concentrates; therefore, the use of plasma-derived factor preparations has been reported to increase the seroprevalence of parvovirus B19 antibodies^29^.

The intravenous injection of the AAV vector induces NAbs against the administered serotype, making the intravenous readministration of the same serotype difficult^30,31^. Strategies for the second administration of the AAV vector should be considered for patients who do not respond to the initial AAV gene therapy or for those who have a diminished therapeutic response. To date, various preclinical studies have been conducted on the treatment of patients with NAbs, *i*.*e*., the administration of immunoglobulin G (IgG)-degrading enzyme, simultaneous administration of empty capsid, plasma exchange, intraportal administration, and use of rapamycin^18,32–35^. The most practical approach is to alter the AAV serotype. Indeed, the readinistration of AAV1 vector reportedly succeeded in monkeys and mice previously treated with AAV5^31^. Recently, the long-term observation of hemophilia A dogs treated with AAV vector suggested NAbs elicited by the vector administration was little cross-reactivity against other serotypes^36^. On the other hand, a report of human clinical trials treated with AAV2 indicated the long-term persistence of NAbs reacted with AAV5 and AAV8^37^. However, we cannot rule out the possibility that the patients already possessed NAbs against other serotypes before the trial, because the seropositivity against other serotypes was not assessed at baseline. Clinical trials to explore whether NAbs for other AAV serotypes are produced after vector administration are also mandated for the readministration.

This study has several limitations. First, we discussed the prevalence of NAbs and time course compared with our previous reports, but we did not assess the same participants over time. The most facilities in previous study^8^ were included in the present study; the previous study incorporated 6 facilities and 5 of them participated in this study (total 14 facilities in this study). We suspected that many previous participants were overlapped, but unable to track which patients have been enrolled in the earlier study. Second, the variation of methodology in the previous studies of NAb prevalence must be considered. In some studies, total antibody binding with AAV capsid was measured, while others employed cell-based neutralizing assay. We used a cell-based assay because total antibody assays detect the binding antibody without any inhibitory effects^2,7,12^. On the other hand, the cell-based assay depends on the cells used in the experiments and the multiplicity of infection (MOI) of AAV^38^. Assessment of neutralizing factors are known to be less sensitive than detecting binding antibodies^4,7,12^. Accordingly, comparison among different studies should be interpreted with caution. Indeed, the global study to measure the binding antibody against AAV5 showed that the higher prevalence in Japanese (29.8%)^9^, compared with those in our study. Furthermore, the interpretation of the prevalence especially in African American population might be overestimated, because the sera had little background information and the number of specimens is limited. Hence, our comparative data with US population should be interpretated with caution. Finally, the NAb titer to clinically suppress the AAV vector transduction depends on the serotype and dose of the AAV vector to be administered^39^. Hence, it is imperative to consider the threshold to judge “NAb-positive” for each drug to be administered.

In conclusion, this study indicates that many patients would benefit from the AAV-mediated gene therapy in Japan. In addition, the seroprevalence is significantly higher in 60 years or older age, and the population has high NAb titers cross-susceptibility to multiple AAVs. Nevertheless, further data on the natural route of AAV infection in the environment, changes in NAb titers after the administration of AAV vectors, and cross-reactivity to other serotypes are warranted.

## MATERIALS AND METHODS

### Study protocol and participants

The Institutional Review Board at Jichi Medical University approved the study protocols (permission number: A19-108), and we obtained written informed consent from all the participants. The study was registered in UMIN-CTR (UMIN-CTR: UMIN000039069). We enrolled 100 healthy volunteers (10 each from males and females in their 20s, 30s, 40s, 50s, and 60s) and consecutive hemophiliacs from Jan 2020 to Mar 2021. The eligibility criteria for hemophiliacs were hemophilia, Japanese-origin man, and age ≥10 years. The exclusion criteria were as follows: 1) Participants unable to give written consent; 2) Participants being treated with antibody drugs including emicizumab; 3) Participants being treated with systemic (oral or injectable) immunosuppression; 4) Participants with possible immune abnormalities such as inflammatory diseases; 5) Participants deemed ineligible by the principal investigator. To minimize the possibility of nonspecific antibody reactions to the AAV vector, patients treated with antibody drugs, including emicizumab, were excluded.

The cross-sectional study was conducted to investigate the seroprevalence of anti-AAV NAbs in hemophiliacs in Japan. The number of samples required for the analysis was first estimated to be 94.65, with a margin of 10% error to analyze the parent target population (6455 hemophiliacs in Japan, the National Survey of Hemostatic Disorders^40^). Therefore, we originally aimed to enroll each of 100 hemophiliacs and healthy volunteers for the study. After enrollment, the target number of hemophilia cases was changed to 200 because the number of patients collaborating in the study was higher than expected during the enrollment period. This change improved the error margin to 7%, further increasing the accuracy of the study results. Healthy volunteers were recruited only from one facility (Jichi Medical University), while hemophiliacs were recruited from 14 facilities in Japan. Each facility enrolled 4–36 hemophilia patients.

### Variables

For healthy participants, information was obtained on age and sex. For hemophiliacs, information on age, type and severity of hemophilia, history of viral infection, and coagulation factor preparation used was obtained. After obtaining informed consent, 10 mL of whole blood was collected by venipuncture with vacuum blood collection tubes (VENOJECT^®^II, TEROMO Corp., Tokyo, Japan) and a 22G needle. The tubes were allowed to stand for 30 min at room temperature, and serum was obtained by centrifugation at 2,000 *g* for 10 min. Then, the serum was aliquoted in 1 mL and stored at −80°C until analysis. The primary endpoint of this study is to determine the percentage of seroprevalence, and to enumerate the gross number of hemophiliacs who will benefit from AAV-mediated gene therapy in Japan. The secondary endpoints were: (1) the correlation of age, sex, viral infection, and types of hemophilia with the seroprevalence of NAb against AAV; and (ii) the establishment of anti-AAV NAb assay.

Normal human sera (aged 19–29 years) from US Caucasians (5 male and female) and US African–Americans (5 male and female) were purchased from Precision for Medicine (Norton, MA).

### AAV vector construction

We introduced a DNA cassette comprising the luciferase gene or the secreted type of the NanoLuc gene driven by the CAG promoter and SV40 polyA between internal terminal repeats of pAAV-CMV (Takara Bio, Shiga, Japan). pRC1, 2, 5, and 6 (for the production of AAV1, 2, 5, and 6, respectively) were purchased from Takara Bio. The capsid sequences of AAV8, AAV9, and AAV3B were synthesized in GenScript Japan (Tokyo Japan) and then cloned into the pRC plasmid. The plasmids for the production of AAV7 (#112863) and AAVrh10 (#112866) were obtained from Addgene (Watertown, MA). Next, the AAV vector was produced by the transfection of AAVpro293 cells (Takara Bio) with three plasmid transfections (pAAV, pRC, and pHelper; Helper-free system). The AAV vector was isolated by CsCl-based ultracentrifugation, as described previously^41^. We confirmed that our AAV vector contained >80% of full particles by analytical ultracentrifugation (data not shown). Finally, the titer of the AAV vector was determined, as described previously^42^.

### Measurement of anti-AAV NAb

The sera from the subjects were heat-inactivated (56°C, 30 min) and diluted with fetal bovine serum (FBS; Thermo Fisher Scientific, Waltham, MA) to 1:1–1:3,000, and then incubated with an AAV vector 1:1 for 1 h at 37°C. Then, the mixture was added to a 96-well plate seeded with Huh-7 cells in triplicate at MOI 1500. We essentially used luciferase as a reporter gene to measure AAV transduction. As the transduction efficacy of Huh-7 with AAV7, AAV9, and AAVrh10 expressing luciferase was lower than the target relative light unit (RLU = 10,000) in the preliminary experiment (data not shown), we used the secreted type of NanoLuc (secNanoLuc) for transduction with AAV7, AAV9, and rh10. Moreover, MOI 150 was used in the case with AAV6 because high transduction efficacy might inhibit the detection of NAb. After 48-h incubation, cells were lysed with 50 μL of 1× Passive Lysis Buffer (Promega, Madison, WI) for the luciferase assay and then stored in a deep freezer. To measure the secNanoLuc activity, 80 μL of the supernatant was collected and mixed with 20 μL of 5× Passive Lysis Buffer (Promega) and stored in a deep freezer. After thawing frozen samples at room temperature, 10 μL of lysates or supernatants were added to a 96-well plate (Berthold Technologies, Bad Wildbad, Germany). The 96-well plate was placed on a luminometer (Centro LB 960, Berthold Technologies), and 50 μL of the Luciferase Assay Reagent (Promega) or Nano-Glo^®^ Luciferase Assay Reagent (Promega) was injected into each well using an automatic injector. The delay and measurement times were set for 2 and 10 s, respectively. All measurements were done at room temperature and completed within 15 min. First, all samples were screened at the dilution of 1:1, and positive samples were selected for further experiments to determine the NAb titer. The serum that inhibited >50% vector transduction was considered a positive sample. The sera of positive samples were diluted with FBS to 1:3, 1:10, 1:30, 1:100, 1:300, 1:1000, and 1:3000, and then the inhibition of vector transduction was assessed. Next, we estimated antibody titers that neutralized 50% vector transduction (ND_50_) by nonlinear regression (inhibitor vs. normalized response, variable slope) using GraphPad Prism 9 (GraphPad, San Diego, CA). We considered 1:1 as ND_50_ of positive samples not to neutralize the vector transduction at 1:3 dilution. On the contrary, ND_50_ to neutralize >50% vector transduction at 1:3000 dilution considered as 1:3000. We employed 1:1 as the cutoff value for NAb-positive in this study because our previous data showed that a low dose of AAV8 vector (5 × 10^11^ vg/kg) was inhibited in mice whose serum NAb titer was 1:1^42^. We consistently used human immunoglobulin (GAMMAGARD^®^, Shire plc, Dublin, Ireland) as a positive control and confirmed that the variation of ND_50_ in each measurement was within 3-fold. We compared this method with the method from our previous study^8^ and confirmed that ND_50_ of human immunoglobulin against AAV5 and AAV8 assessed by either of the methods was not statistically different (*P* = 0.1347 and *P* = 0.3432, respectively).

### Statistical analysis

All statistical analyses were performed with GraphPad Prism 9 or JMPpro 16 (SAS Institute, Cary, NC). The categorical variables are presented as percentages. The continuous variables, *e*.*g*., age and NAb titers, are presented with each data point in figures. The normality of the distribution of continuous variables was examined using the Kolmogorov–Smirnov test. As the titers of NAb were not normally distributed (data not shown), we performed non-parametric testing for the comparison. Between-group differences in frequencies were examined with the Fisher’s extract test, while differences in frequencies among ≥3 groups were examined with the *χ*^2^ test for trend. In addition, values between the two groups were compared with the Mann–Whitney *U* test. The differences between two groups containing >3 groups were determined by the Friedman test (when each row was matched) or the Kruskal–Wallis test (when each row was not matched) with the posthoc multiple comparison test. Posthoc pairwise comparison between the groups (*i*.*e*., patients vs. healthy volunteers) among >3 groups (*i*.*e*., age groups) was analyzed using the two-way analysis of variance with a posthoc multiple comparison test. The correlation coefficient was measured using Spearman’s rank correlation coefficient. We applied factor analysis to identify latent factors to explain the variance. Furthermore, the logistic regression analysis was used to examine the correlation of independent variables with one dichotomous dependent variable. We did not obtain data on HCV, HIV, and HBV in three, three, and six cases, respectively. The patients with missing data were excluded from the comparison between those with and without viral infection. In contrast, we included all participants in the multivariate analysis. In this study, *P* < 0.05 was considered statistically significant. Besides, we created the topological phylogenetic tree using the maximum likelihood method with Molecular Evolutionary Genetics Analysis 11 software (https://www.megasoftware.net/home).

## Data Availability

All data produced in the present study are available upon reasonable request to the authors.

## ACKNOWLEDGMENTS

This work was supported by AMED [JP18pc0101030]. Optima XE-90 were subsidized by JKA through its promotion funds from KEIRIN RACE. We thank Sachiyo Kamimura, Mika Kishimoto, Yaeko Suto, Tamaki Aoki, Mai Hayashi, Yuiko Ogihara, Nagako Sekiya, Noguchi Tomoko, Hiromi Ozaki, and Hiroko Hayakawa of Jichi Medical University for their technical assistance. Moreover, we gratefully appreciate the healthy volunteers and the hemophiliacs who participated in this study. The authors would like to thank Enago (www.enago.jp) for the English language review.

## AUTHOR CONTRIBUTIONS

Y.K. conducted the experiments, analyzed the data, and wrote and revised the manuscript. N.B., R.W. and T.H. conducted the experiments, analyzed the data, and revised the manuscript. S.Y. designed the study, conducted the experiments, and revised the manuscript. A.N., K.A., N.S., T.M., A.S., S.H. N.Y., T.F., T.O., H.T., M.T., T.M., J.Y., M.S., M.N., Y.Y., K.Y., and K.N. recruited the study participants and obtained serum and data from the participants. M.H., N.K., A.K., H.M., and Y.S. analyzed the data and revised the manuscript. S.I. supervised all statistical analysis and revised the manuscript. T.O. designed the study, recruited the study participants, collected blood samples from all healthy volunteers, analyzed the data, and wrote and revised the manuscript. All authors approved the final version of the manuscript.

## DECLARATION OF INTERESTS

All authors have no competing financial interests to declare.

### Data Availability Statement

The data that support the findings of this study are available from the corresponding author, [T.O.], upon reasonable request.

