## Supplemental Table S1, S2, Figure S1, S2, and S3 for "The seroprevalence of neutralizing antibodies against the adeno-associated virus capsids in Japanese hemophiliacs": Supplemental Files_R1_final.docx

**Table S1.** Comparison of AAV Nab positive subjects for each serotype among generations

|  | Healthy volunteers |  |
| --- | --- | --- |
| Serotype | Chi-square, df | *P* |
| AAV1 | 15.41, 1 | <0.0001 |
| AAV2 | 14.61, 1 | 0.0001 |
| AAV3B | 11.76, 1 | 0.0006 |
| AAV5 | 10.19, 1 | 0.0014 |
| AAV6 | 18.84, 1 | <0.0001 |
| AAV7 | 14.10, 1 | 0.0002 |
| AAV8 | 6.556, 1 | 0.0105 |
| AAV9 | 11.28, 1 | 0.0008 |
| AAVrh10 | 7.922, 1 | 0.0049 |
|  | Patients |  |
| Serotype | Chi-square, df | *P* |
| AAV1 | 27.11, 1 | <0.0001 |
| AAV2 | 30.06, 1 | <0.0001 |
| AAV3B | 29.36, 1 | <0.0001 |
| AAV5 | 24.66, 1 | <0.0001 |
| AAV6 | 30.47, 1 | <0.0001 |
| AAV7 | 30.19, 1 | <0.0001 |
| AAV8 | 19.75, 1 | <0.0001 |
| AAV9 | 28.65, 1 | <0.0001 |
| AAVrh10 | 24.46, 1 | <0.0001 |

AAV, adeno-associated virus; CI, confidence interval; Nab, neutralizing antibody

**Table S2.** Comparison of AAV Nab titer between patients and healthy volunteers according to generations

| AAV1 | Generation | Predicted mean difference (95% CI) | *P* |
| --- | --- | --- | --- |
|  | 10–20s | −66.2 (−284.6 to 152.2) | 0.9419 |
|  | 30s | 37.61 (−195.5 to 270.8) | 0.9965 |
|  | 40s | −41.62 (−269.2 to 186.0) | 0.9937 |
|  | 50s | −46.61 (−276.8 to 183.6) | 0.9899 |
|  | 60s– | −72.94 (−310.8 to 164.9) | 0.9391 |
| AAV2 | Generation | Predicted mean difference (95% CI) | *P* |
|  | 10–20s | −38.49 (−223.9 to 147.0) | 0.9887 |
|  | 30s | 23.84 (−174.2 to 221.8) | 0.9991 |
|  | 40s | −31.66 (−224.9 to 161.6) | 0.9962 |
|  | 50s | −82.82 (−278.3 to 112.7) | 0.7987 |
|  | 60s– | −82.93 (−284.9 to 119.0) | 0.8188 |
| AAV3B | Generation | Predicted mean difference (95% CI) | *P* |
|  | 10–20s | −80.23 (−379.8 to 219.4) | 0.9653 |
|  | 30s | 61.41 (−258.4 to 381.3) | 0.9921 |
|  | 40s | −82.01 (−394.3 to 230.2) | 0.968 |
|  | 50s | −55.21 (−371.0 to 260.6) | 0.9949 |
|  | 60s– | −197.6 (−523.9 to 128.7) | 0.4677 |
| AAV5 | Generation | Predicted mean difference (95% CI) | *P* |
|  | 10–20s | −14.45 (−62.75 to 33.84) | 0.9449 |
|  | 30s | 10.15 (−41.41 to 61.72) | 0.9911 |
|  | 40s | −4.685 (−55.02 to 45.65) | 0.9998 |
|  | 50s | −8.222 (−59.13 to 42.69) | 0.9965 |
|  | 60s– | −11.42 (−64.02 to 41.18) | 0.9861 |
| AAV6 | Generation | Predicted mean difference (95% CI) | *P* |
|  | 10–20s | −67.99 (−319.2 to 183.2) | 0.9637 |
|  | 30s | 16.95 (−251.3 to 285.2) | >0.9999 |
|  | 40s | −67.36 (−329.2 to 194.5) | 0.9707 |
|  | 50s | −76.1 (−340.9 to 188.7) | 0.9533 |
|  | 60s– | −45.16 (−318.8 to 228.4) | 0.9961 |
| AAV7 | Generation | Predicted mean difference (95% CI) | *P* |
|  | 10–20s | −65.18 (−277.3 to 146.9) | 0.9386 |
|  | 30s | 32.48 (−194.0 to 259.0) | 0.998 |
|  | 40s | −9.235 (−230.3 to 211.9) | >0.9999 |
|  | 50s | −82 (−305.6 to 141.6), | 0.8784 |
|  | 60s– | −72.88 (−303.9 to 158.1) | 0.9318 |
| AAV8 | Generation | Predicted mean difference (95% CI) | *P* |
|  | 10–20s | −28.95 (−171.1 to 113.2) | 0.9896 |
|  | 30s | 7.659 (−144.1 to 159.4) | >0.9999 |
|  | 40s | −16.36 (−164.5 to 131.8) | 0.9994 |
|  | 50s | −50.32 (−200.1 to 99.47) | 0.9126 |
|  | 60s– | −161.4 (−316.1 to −6.587) | 0.0366 |
| AAV9 | Generation | Predicted mean difference (95% CI) | *P* |
|  | 10–20s | −28.37 (−160.9 to 104.1) | 0.987 |
|  | 30s | 20.91 (−120.5 to 162.4) | 0.9977 |
|  | 40s | −13.85 (−151.9 to 124.2) | 0.9996 |
|  | 50s | −19.51 (−159.2 to 120.1) | 0.9982 |
|  | 60s– | −82.07 (−226.4 to 62.22) | 0.5365 |
| AAVrh10 | Generation | Predicted mean difference (95% CI) | *P* |
|  | 10–20s | −62.11(−270.9 to 146.7) | 0.9462 |
|  | 30s | 30.99 (−191.9 to 253.9) | 0.9983 |
|  | 40s | −51.97 (−269.6 to 165.7) | 0.9788 |
|  | 50s | −55.04 (−275.1 to 165.1) | 0.9741 |
|  | 60s– | −112.7 (−340.0 to 114.7) | 0.6751 |

AAV, adeno-associated virus; CI, confidence interval; Nab, neutralizing antibody

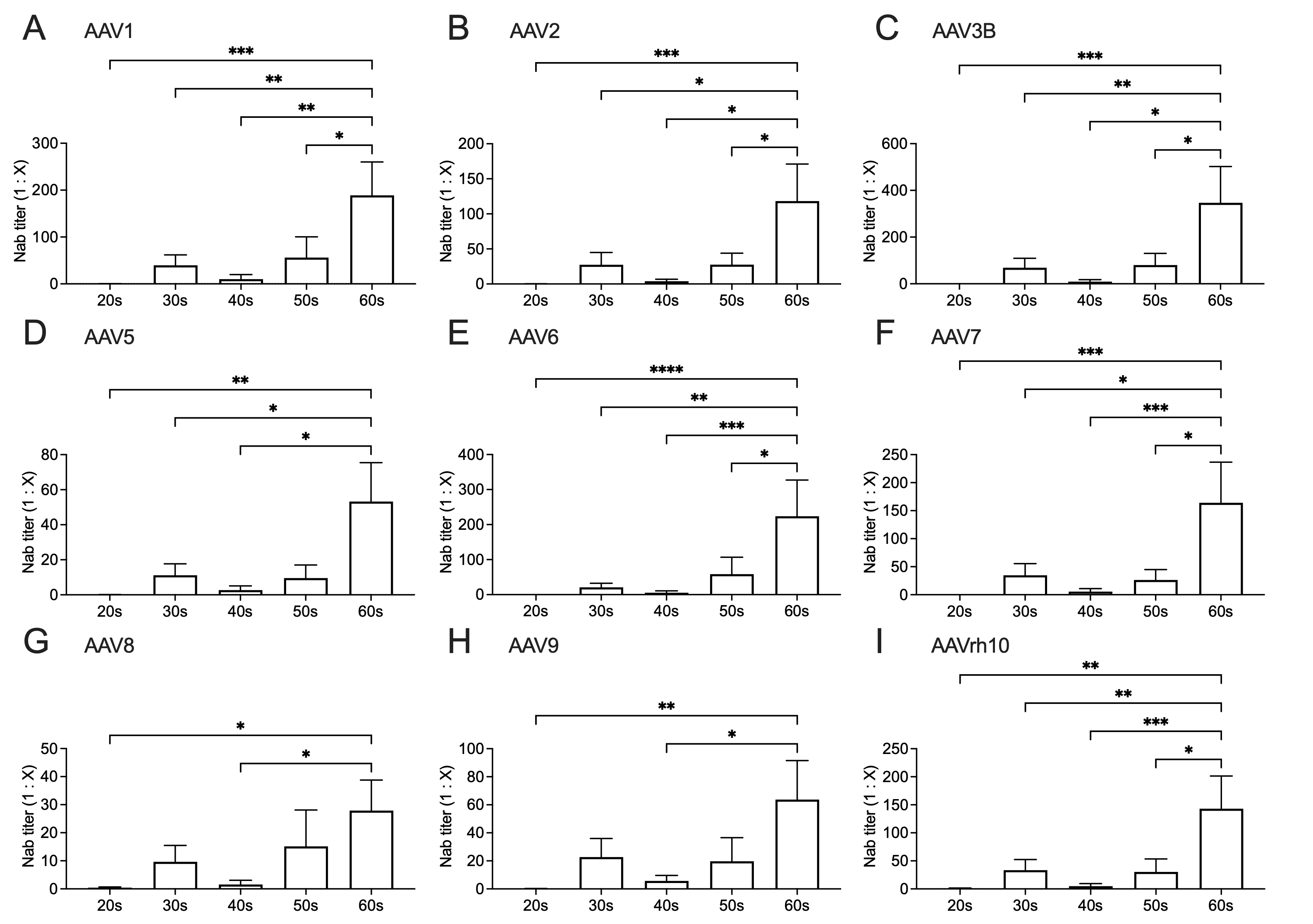

**Figure S1. Comparison of the AAV Nab titer among age generations in healthy volunteers.** The Nab titer to each AAV serotype (A: AAV1, B: AAV2, C: AAV3B, D: AAV5, E: AAV6, F: AAV7, G: AAV8, H: AAV9, I: AAVth10) in each age generation in healthy volunteers. Values are mean ± SEM (n = 20). The value comparison between the groups is analyzed by the Kruskal–Wallis test with post hoc multiple comparisons. **P* < 0.05, ***P* < 0.01, and ****P* < 0.001. AAV, adeno-associated virus; Nabs, neutralizing antibodies.

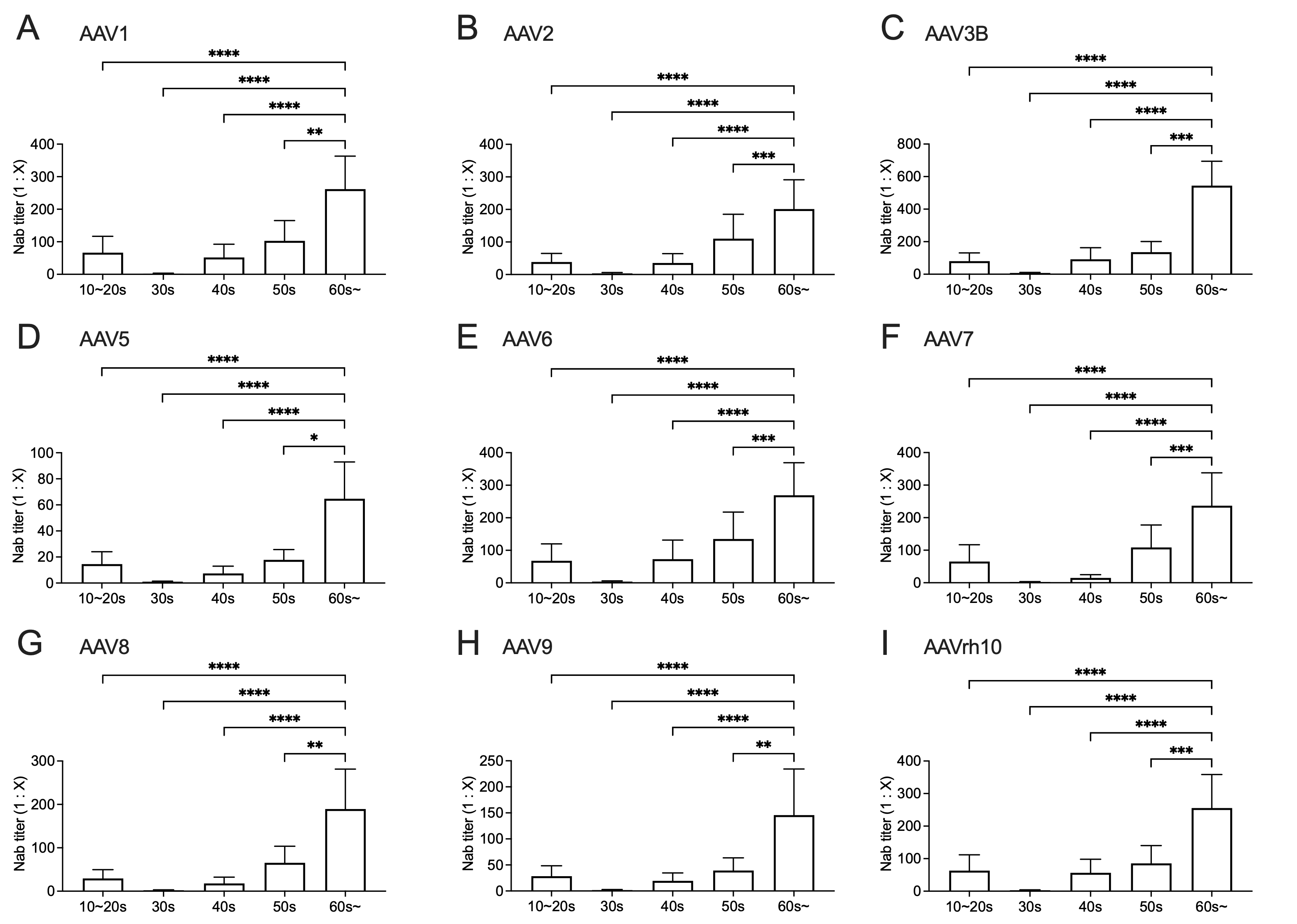

**Figure S2. Comparison of the AAV Nab titer among age generations in hemophilia patients.** The Nab titer to each AAV serotype (A: AAV1, B: AAV2, C: AAV3B, D: AAV5, E: AAV6, F: AAV7, G: AAV8, H: AAV9, I: AAVth10) in each age generation in hemophiliacs. Values are mean ± SEM (10–20s, n = 59; 30s, n = 38; 40s, n = 44; 50s, n = 41; 60s–, n = 34;). The value comparison between the groups is analyzed by the Kruskal–Wallis test with posthoc multiple comparisons. **P* < 0.05, ***P* < 0.01, ****P* < 0.001, and *****P* < 0.0001. AAV, adeno-associated virus; Nabs, neutralizing antibodies.

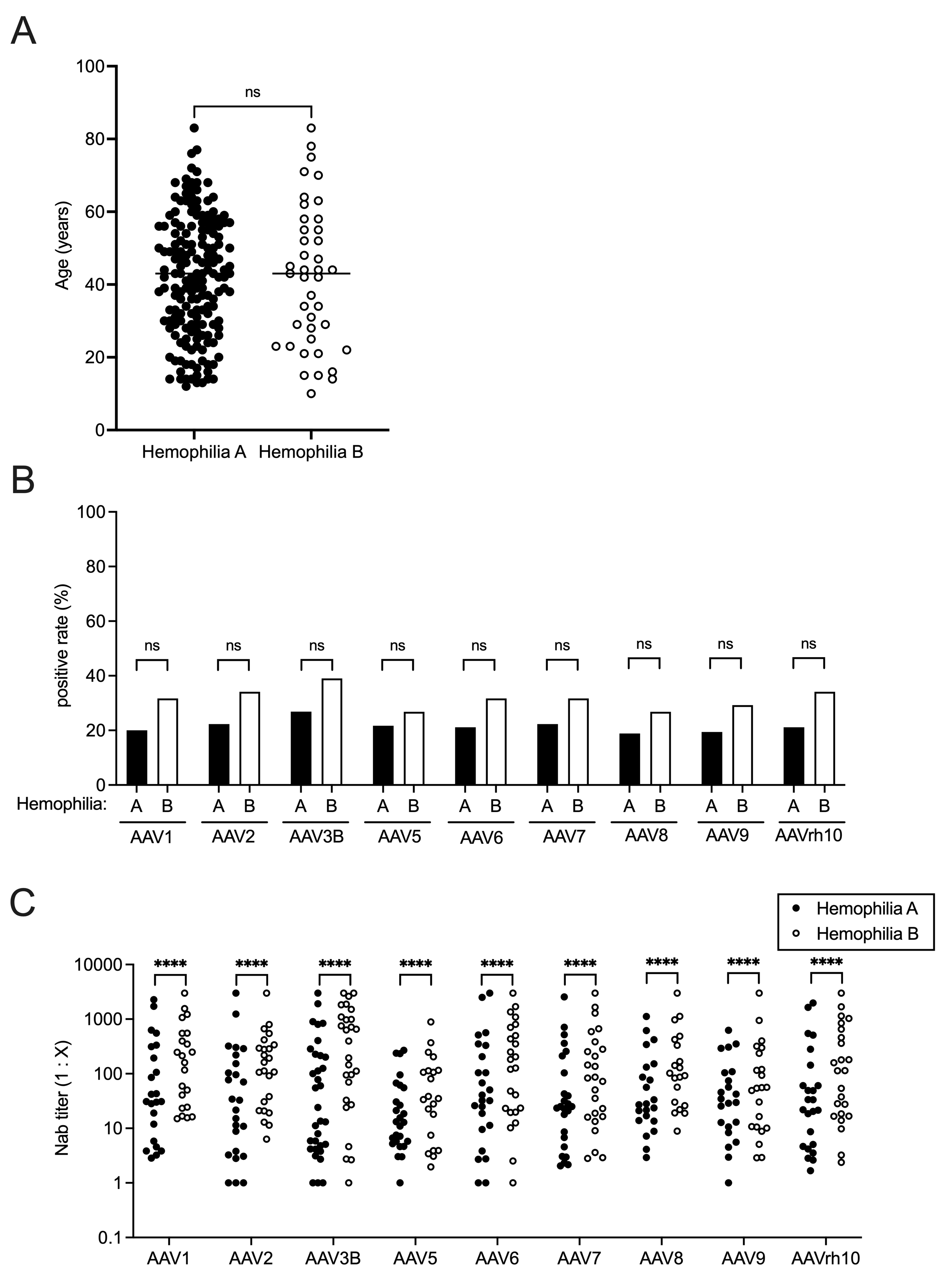

**Figure S3. Comparison of the seroprevalence and titer of AAV Nabs between hemophilia A and hemophilia B.** (A) Comparison of the ages between the groups. The statistical difference was analyzed by the Mann–Whitney *U* test. Bar, median. (B) The seroprevalence of AAV Nabs. The between-group statistical significance was analyzed by the Fisher’s extract test. (C) The Nab titer against AAV. The value comparison between the groups is analyzed by the Mann–Whitney *U* test. *****P* < 0.0001. AAV, adeno-associated virus; Nabs, neutralizing antibodies; n.s., not significant.
